# Establishment of *in silico* prediction of adjuvant chemotherapy response from active mitotic gene signature in non-small cell lung cancer

**DOI:** 10.1101/2025.03.14.25322930

**Authors:** Eun-Ji Kwon, Hee Sang Hwang, Eunhyong Chang, Joon-Yong An, Hyuk-Jin Cha

**Author notes:** To whom correspondence should be addressed to, Prof. Hyuk-Jin Cha, Ph.D., College of Pharmacy, Seoul National University, 1 Gwanak-ro, Gwanak-gu, Seoul 08826, Republic of Korea.

## Abstract

Conventional chemotherapeutics exploit cancer’s hallmark of active cell cycling, primarily targeting mitotic cells. Consequently, the mitotic index (MI), representing the proportion of cells in mitosis, serves as both a prognostic biomarker for cancer progression and a predictive marker for chemo-responsiveness. In this study, we developed a transcriptome signature to predict the chemotherapeutic responsiveness based on the Active Mitosis Signature Enrichment Score (AMSES), a computational metric previously established to estimate the active mitosis using multi-omics data from The Cancer Genome Atlas (TCGA) lung cancer cohorts, lung adenocarcinoma (LUAD) and lung squamous cell carcinoma (LUSC) patients. Leveraging advanced machine learning techniques, we enhanced the predictive power of AMSES and developed ‘AMSES for chemo-responsiveness’, termed A4CR. Comparative analysis revealed a strong correlation between A4CR and the MI of 69 cases from separated non-small cell lung cancer (NSCLC) cohort. The utility of A4CR as a therapeutic biomarker was validated through *in silico* analysis of public datasets, encompassing transcriptomic profiles of cancer cell lines (CCLs) and their corresponding multiple drug response data as well as clinicogenomic data from TCGA. These findings highlight the potential of integrating gene signatures with machine learning and large-scale datasets to advance precision oncology and improve therapeutic decision-making for cancer patients.

## Introduction

Since the initial definition of the hallmarks of cancer in 2000 (1), additional hallmarks have been identified and redefined approximately every decade (2,3). These evolving frameworks have provided promising targets and approaches for addressing the diverse and individualized nature of cancer, leading to novel therapeutic strategies such as immune checkpoint inhibitors and targeted therapies. Nonetheless, conventional chemotherapeutics targeting actively proliferating cells—one of cancer’s most fundamental features—remain crucial as either first-line treatments or last-resort options (4). However, as well as their substantial toxic side effects on renewing organs, these agents pose limited clinical benefit for some cancer subtypes due to acquired chemoresistance or slower growth rate (5–7).

To overcome these challenges, histopathological biomarkers such as the mitotic index (MI) and Ki-67 labeling index, indicative of tumor growth rate, grade, and patient survival (8,9), have been widely utilized to predict chemotherapy benefit (10,11). Proliferative activity, often measured through histone H3 phosphorylation at serine 10, requires labor-intensive evaluations by pathologists (12). However, since mitosis is governed by a complex regulatory network involving structural proteins, molecular motors, kinases, and phosphatases (9), a more integrative approach is necessary. Rather than relying on single-protein immunohistochemistry, evaluating the entire mitotic apparatus network provides a more comprehensive assessment of mitotic activity (13).

In this context, transcriptomic signatures extend beyond diagnostic and prognostic markers (14), serving as phenotypic indicators of treatment response and resistance (15,16). Scoring mitosis-related genes has been proposed, including a 44-gene signature linked to early metastasis (17) and a 54-gene set associated with poor immunotherapy response (18). Additionally, an analysis of over 1,000 periodic genes identified a “mitotic trait” that classifies tumors based on cell cycle stage, correlating with genetic alterations, tumor subtypes, and patient survival, while pinpointing 67 core genes essential for cell cycle progression (19). Recent studies underscore the predictive power of gene signatures in identifying druggable targets and repositioning existing drugs (20–22). Integrative approaches incorporating advanced machine learning frameworks are paving the way for precision oncology and personalized therapeutic strategies addressing cancer’s complexity and heterogeneity (23,24).

In this study, we aimed to develop a transcriptome signature to represent the mitotic index and serve as a robust predictive marker for chemotherapeutic responsiveness. Using the machine learning-based Active Mitosis Signature Enrichment Score (AMSES), validated in a cohort of 69 Korean non-small cell lung cancer (NSCLC) patients, we established ‘AMSES for chemo-responsiveness’ (A4CR). The predictive capacity of A4CR was further validated using The Cancer Genome Atlas (TCGA) lung cancer cohorts and multiple drug response datasets from cancer cell lines. Given the significant side effects of traditional chemotherapeutics, this simple approach for predicting chemo-responsiveness offers a valuable tool for clinical decision-making, balancing the risk-benefit of conventional chemotherapies.

## Material and Methods

### Recurrence Analysis

We investigated the difference of recurrence patterns of both Korean NSCLC and TCGA (TCGA-LUAD and TCGA-LUSC) cohorts according to the history of adjuvant chemotherapy. Patients were classified into tertiles according to mitosis count or geneset score values or gene expression or protein phosphorylation, with the lowest 33% designated as the “low” group and the highest 33% as the “high” group. Survival analysis was conducted to compare recurrence rates between the low and high groups. The hazard ratio (HR) and P-value were calculated using Cox proportional hazards regression and the log-rank test, implemented with the R package ‘survival’ (v3.4-0).

### Correlation between Drug Response and Recurrence Rate

Using data from TCGA LUAD and LUSC, cohorts, we classified patients who received chemotherapy according to their “Treatment_best_response” values: patients labeled “CR” (complete response) and “PR” (partial response) were categorized as “Response,” while those labeled “PD” (progressive disease) and “SD” (stable disease) were grouped as “Disease.” Subsequently, a hypergeometric test was used to evaluate the association between drug response and recurrence status.

### Differential Expression Analysis and Functional Enrichment

To identify differentially expressed genes (DEGs) between recurrence and non-recurrence groups, Wald tests were conducted using the R package DESeq2 (v3.15). DEGs were selected based on an FDR-adjusted P-value of less than 0.05 and an absolute fold-change greater than 2. Geneset enrichment analysis (GSEA) was then performed to identify pathways significantly represented at the top or bottom of ranked gene lists from differential expression analysis between recurrence and non-recurrence groups. The GSEA was conducted using the R package fgsea, with parameters set to minSize=10, maxSize=500, and nperm=100,000.

### Collection of Cancer Cell Line and Drug response Datasets

Human CCL datasets were obtained from the DepMap portal (https://depmap.org/portal/, version 24Q2), containing transcriptome and drug response data for over 1,000 CCLs across more than 30 cancer types. Drug response data were sourced from the CTRP (25), PRISM (26), and GDSC (27) databases. The area under the dose-response viability curve (AUC) was used as a measure of drug sensitivity, with a lower AUC indicating higher sensitivity.

### Correlation Between Geneset Scores and Drug Response in Cancer Cell Lines

Geneset scores for Hallmark gene sets from the MSigDB database were computed for individual samples in the CCLE dataset using Gene Set Variation Analysis (GSVA) applied to TPM expression values. For CCLs overlapping with the CTRP dataset, correlations between gene set scores and drug response (AUC values) were analyzed. Positive correlation coefficients indicate that higher gene set enrichment is associated with higher AUC values (lower drug sensitivity), while negative coefficients indicate greater sensitivity.

### Comparative Analysis of Drug Sensitivity Based on Geneset Enrichment Scores in Cancer Cell Lines

Optimized geneset (A4CR, RF and Ridge), AMSES geneset scores for individual samples in the CCLE and GDSC datasets were calculated using GSVA on Transcripts Per Million (TPM) values for all genes, via the R package gsva. Additionally, cell lines were divided into high-score and low-score groups based on the median geneset score. A series of t-tests was performed to compare the average AUC distributions of 14 chemotherapy drugs between these groups, leveraging data from the CTRP, PRISM, and GDSC datasets.

### Optimization of the AMSES Geneset for Mitotic Index and Recurrence Status

The AMSES geneset was optimized to better represent the Mitotic Index (MI) value and recurrence status (Recur status). A total of 69 patients were included in the analysis. Patients were categorized into two groups based on the median MI value (“H” for high and “L” for low) and into two groups based on recurrence status (“R” for recurrent and “NR” for non-recurrent).

### Identification of Genes Associated with Mitotic Index

1. Correlation Analysis: Pearson correlation coefficients between the 126 genes in the AMSES geneset and the actual MI values were calculated to evaluate their individual associations.
2. Random Forest Model: A Random Forest model (RandomForestClassifier from the scikit-learn package)was implemented to classify patients into the “H” and “L” MI groups using the following hyperparameters: max_depth: 10, max_features: ‘sqrt’, min_samples_leaf: 1, min_samples_split: 3, n_estimators: 93. Feature importance values for each gene were derived from the model.
3. Ridge Logistic Regression Model (LogisticRegression from the scikit-learn package): A Ridge-penalized logistic regression model was developed to classify patients into “H” and “L” groups with the following hyperparameters: solver: ‘saga’, C: 3.755, max_iter: 100. Coefficient values for each gene were extracted to assess their contribution to the prediction.

### Identification of Genes Associated with Recurrence Status

1. Random Forest Model: A Random Forest model was constructed to classify patients into “R” and “NR” groups based on recurrence status, using the following hyperparameters: max_depth: 30, max_features: ‘sqrt’, min_samples_leaf: 1, min_samples_split: 8, n_estimators: 192. Feature importance values for each gene were obtained.
2. Ridge Logistic Regression Model: A Ridge-penalized logistic regression model was developed to classify patients into “R” and “NR” groups with the following hyperparameters: solver: ‘saga’, C: 1.570, max_iter: 1000. Coefficient values for each gene were calculated to determine their predictive utility.

### Model Implementation and Evaluation

All models were implemented using the scikit-learn (sklearn) module. Optimal hyperparameters were identified using RandomizedSearchCV. Model performance was evaluated using the following metrics: Accuracy, Precision, Recall, F1-score and AUC-ROC. These metrics ensured robust validation of the models in predicting both MI values and recurrence status.

### Gene Set Construction and Recurrence Analysis Using GSVA Scores

Genes were ranked by feature importance and coefficient values, and 126 cumulative gene sets were created by progressively adding one gene at a time. GSVA scores were calculated using TPM values for each gene set. Patients were categorized into “High” and “Low” groups based on the upper and lower 33% of GSVA scores. Recurrence risk was evaluated for each gene set using Cox proportional hazards regression and the log-rank test in the R package survival (v3.4-0). The gene set with the lowest hazard ratio was identified as the most prognostically significant.

### Evaluation of Predictive Performance Using ROC Analysis

To assess the predictive performance of the optimal gene sets constructed for each model in predicting Mitotic index values, we categorized patients into groups based on the lowest tertile of Mitotic index values. Receiver Operating Characteristic (ROC) analysis was then performed for each gene set. The area under the ROC curve (AUROC) was used as a quantitative metric to evaluate the predictive accuracy of each method, with higher AUROC values indicating superior predictive performance.

### Acquisition of Gene Expression and Clinical Data from the TCGA Cohort

RNA-sequencing (RNA-seq) data for patients with lung adenocarcinoma (LUAD), lung squamous cell carcinoma (LUSC), breast invasive carcinoma (BRCA), colon adenocarcinoma (COAD), and head and neck squamous cell carcinoma (HNSC) were obtained from the TCGA legacy gene expression dataset using the GDCquery function in the R package TCGAbiolinks. The dataset includes read counts and TPM values for 19,925 protein-coding genes. Corresponding clinical data for individual samples were also retrieved using the GDCquery function in TCGAbiolinks, enabling integrative analyses of gene expression and clinical outcomes.

### Analysis of A4CR Gene Set Scores and Association with Recurrence Risk and Drug Response

For each cohort, A4CR gene set scores were calculated using Gene Set Variation Analysis (GSVA). Patients were then stratified into “High” and “Low” groups based on the upper and lower 33% of A4CR scores, respectively. The recurrence risk for patients in these groups was evaluated using survival analysis. Additionally, to examine the relationship between A4CR scores and chemotherapeutic response, treatment response categories were defined based on the Treatment_best_response values. Patients classified as “Complete Response” or “Partial Response” were grouped as “Response,” while those with “Progressive Disease” or “Stable Disease” were grouped as “Disease.” A hypergeometric test was subsequently performed to evaluate the statistical association between drug response categories and the high or low A4CR score groups.

### Phosphoproteome data analysis

To analyze recurrence risk based on phosphoproteomic data from K-NSCLC (28), patients were stratified into “High” and “Low” groups according to the upper and lower 33% of phosphorylation intensity values for specific substrates. Two substrates were utilized in this analysis: histone H3 encoded by H3F3A and Polo-like kinase 1 encoded by PLK1. For PLK1, the phosphorylation intensity at threonine 210 (Thr210) was used as the defining metric. In the case of H3F3A, phosphorylation intensity at serine 11 (Ser11) was analyzed instead of serine 10 (Ser10), as Ser11 phosphorylation is the relevant modification for this study. Recurrence risk for patients in the High and Low groups was then evaluated using survival analysis.

## Result

### Reduced Recurrence of cancer patients with high mitosis index after chemotherapy

To evaluate the clinical outcome of conventional chemotherapeutics (hereafter chemotherapy), a clinical cohort integrating transcriptome, proteome, therapeutic regimen, and Response Evaluation Criteria in Solid Tumors (RECIST) data is essential. The Korean NSCLC discovery cohort used in this study includes 200 patients with multiomics datasets and medical treatment records (hereafter, K-NSCLC cohort) out of a total of 231 patients (28). However, the K-NSCLC cohort lacks RECIST data. Therefore, as an alternative, we used post-chemotherapy recurrence status as a surrogate marker for drug responsiveness instead of RECIST data. To validate this approach, we first examined the relationship between drug responsiveness and recurrence in patients who experienced tumor relapse (i.e., non-primary tumors), stratifying cases based on whether chemotherapy was administered as adjuvant therapy or post-recurrence treatment. Notably, when chemotherapy was used as an adjuvant therapy, analysis of lung adenocarcinoma (LUAD) and lung squamous cell carcinoma (LUSC) in The Cancer Genome Atlas (TCGA) cohorts (i.e., TCGA-LUAD and TCGA-LUSC) revealed that patients with favorable chemotherapeutic responses in the RECIST classification [i.e., Response: Complete Response (CR) and Partial Response (PR)] exhibited significantly lower recurrence risks compared to those with poor responses [i.e., Disease: Stable Disease (SD) and Progressive Disease (PD)], as determined by Pearson’s Chi-squared test (Fig. S1A) but not after recurrence occurs (Fig. S1B). This finding indicates that recurrence risk after adjuvant chemotherapy could serve as an alternative to RECIST data in subsequent analyses.

To further investigate, the K-NSCLC cohort (28), comprising 231 non-small cell lung cancer (NSCLC) patients with transcriptomic and (phospho)proteomic datasets, was examined. Of the 231 patients, 31 were excluded due to missing clinical records or transcriptomic data (‘No data’). The remaining 200 patients were categorized into subgroups based on their treatment regimens: i) The ‘Chemo’ group (78 patients) includes 36 patients who received chemotherapy alone and 42 patients who initially received chemotherapy but later relapsed, requiring additional treatment with ionizing radiation (IR) and/or targeted therapy. ii) The ‘IR’ group (46 patients) consists of 12 patients who received IR therapy alone. iii) The ‘Target’ group (23 patients) includes 9 patients who received targeted therapy alone. iv) The ‘No therapy’ group (100 patients) comprises patients who did not receive any form of therapy during the observation period (Fig. 1A).

**Figure 1.**
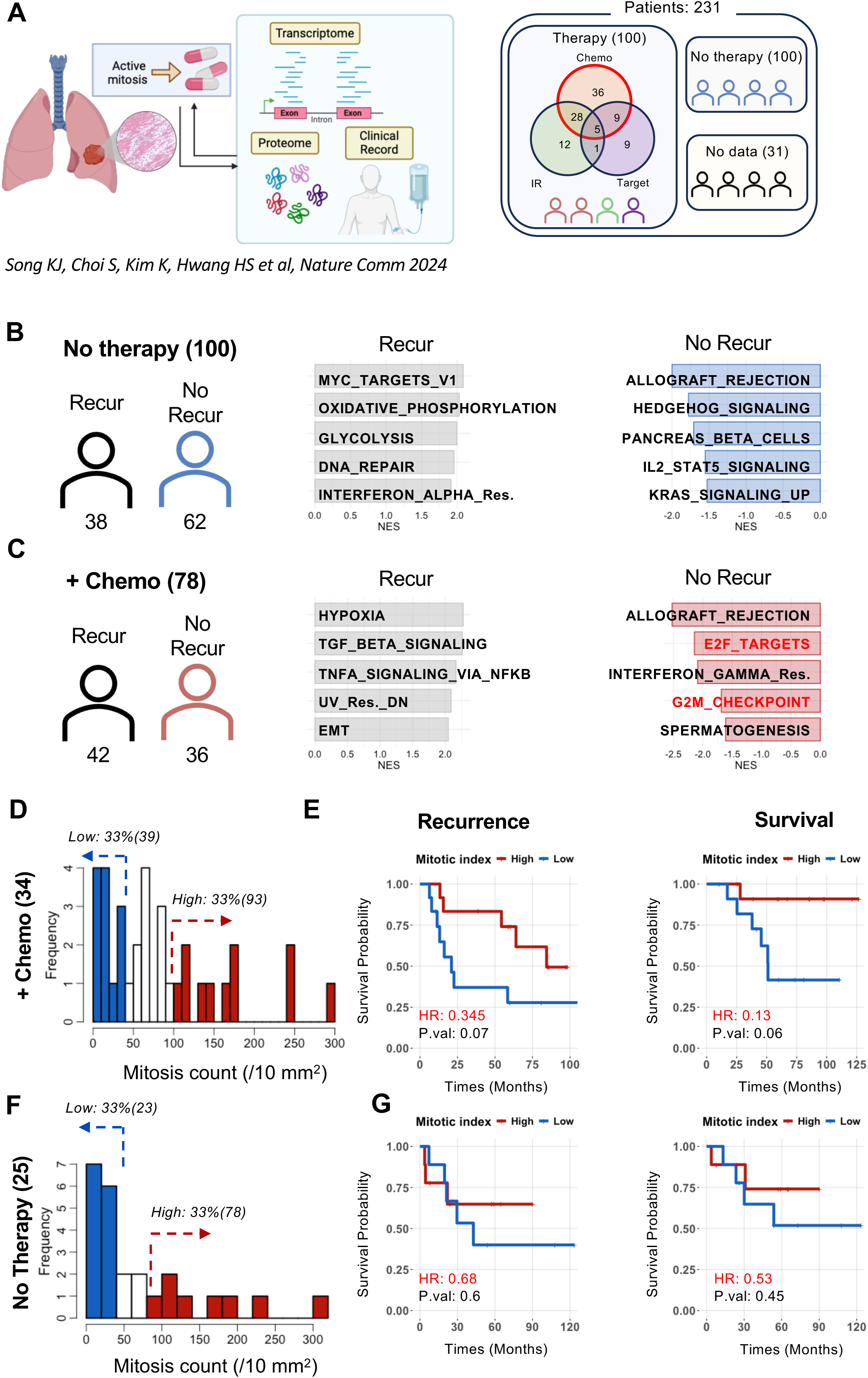
Association of Mitotic Index and Recurrence Risk in Korean Lung Cancer Patients. (A) Schematic illustration of omics data derived from Korean lung cancer patients, as described by Sung et al. (B) Gene Set Enrichment Analysis (GSEA) results comparing patients with and without recurrence within the “No-therapy” group. (C) GSEA results comparing patients with and without recurrence within the “+Chemo” group. (D, F) Distribution of mitotic index values across patient groups, categorized into tertiles: High (top 33%) and Low (bottom 33%). (E, G) Risk analysis of recurrence (left panels) and overall survival (right panels) for patients in the Low and High mitotic count groups.

The chemotherapies and their corresponding modes of action used in the K-NSCLC cohort are listed in Figure S1C. Differentially expressed genes (DEGs) between recurrence and non-recurrence groups in each subgroup (Fig. S1D), were further analyzed using comprehensive Geneset Enrichment Analysis (GSEA). Sixty-two patients with no recurrence (No Recur, Fig. 1B) receiving no therapy, exhibited significant enrichment of genesets associated with ‘signaling pathways’ such as ‘HEGEHOG’,’KRAS’ and ‘IL2_STAT5’ (Fig. 1B). In other hand, thirty-six patients with no recurrence (No Recur, Fig. 1C) in group of ‘Chemo’, showed high enrichment of genesets associated to active cell cycle processes, such as “E2F_TARGETS,” and “G2/M_CHECKPOINT,” (Fig. 1C), while distinct enrichment of typical chemoresistance genesets, including ‘epithelial mesenchymal transition (EMT)’ (22) and ‘TGF beta signaling’, one of main upstream signaling for inducing EMT (29,30). These data suggest that a conventional anti-cancer chemotherapy give a better clinical outcome to the patients with fast-proliferating cancers. Conversely, in the subgroup of patients treated with irradiation alone (‘IR’), the no recurrence group demonstrated an activated “INTERFERON” response (Fig. S1E).

Thus, recurrence status in the K-NSCLC cohort, which closely correlated with chemo-responsiveness, was utilized as a surrogate for RECIST data. To further explore the relationship between active cell cycle dynamics and chemotherapeutic outcomes, a subset of patients with available mitotic index values (MIV, ranging from 0 to 300) was categorized into three groups—‘high’, ‘intermediate’, and ‘low’—based on the number of mitoses, a well-established histopathological marker of active mitosis. Among the 34 patients in the ‘Chemo’ group with available MIV data, patients were subcategorized according to their MIV values (Fig. 1D). Analysis of clinical outcomes, including recurrence and survival, revealed a significant distinction between the high group (MIV > 93) and the low group (MIV < 39). These findings clearly demonstrate that MIV serves as a key determinant of clinical outcomes following chemotherapy. In contrast, a similar analysis of 25 patients in the ‘No Therapy’ group with available MIV data showed no clear association between MIV and clinical outcomes (Figs. 1F and 1G).

### Correlation of mitosis geneset scores to chemo-responsiveness in cell line data

Our findings that mitotic index of cancer patients is closely associated to the clinical outcome of chemotherapy, would be a theoretical analysis for empirical results. In turn, we hypothesized that the chemo-responsiveness of each individual cancer cells, would be predicted by a type of quantitative biomarker to represent the active mitosis. While phosphoproteome data have been generated and multiple datasets are publicly available, the extensive availability of transcriptome datasets has positioned them as a key resource, widely used not only for correlation analyses but also for defining cellular characteristics. To this end, we first conducted a correlation analysis between 50 Hallmark genesets, represented by their enrichment scores, and the responses to FDA-approved drugs, as determined by the area under the dose-response curve (AUC), across 820 cell lines (Fig. 2A). In this analysis framework, a negative correlation coefficient indicates that a high mitotic gene score predicts enhanced chemo-responsiveness. Consistent with the clinical observations in Figure 1, multiple genesets representing active cell cycle processes, including ‘G2M_CHECKPOINT’ and ‘E2F_TARGETS,’ demonstrated significant negative correlations with chemotherapeutic drug sensitivity (Figs. S2A and 2B). In contrast, two genesets, ‘KRAS_SIGNALING_UP’ and ‘ESTROGEN_RESPONSE_EARLY,’ exhibited strong positive and negative correlations with chemotherapy (drugs in red) and targeted therapy (drugs in light blue), respectively (Figs. 2B and S2B). The KRAS_SIGNALING_UP gene set is typically activated in tumors where receptor tyrosine kinases (RTKs), such as epidermal growth factor receptor (EGFR), function as oncogenes (31). Consequently, a significant negative correlation of the KRAS_SIGNALING_UP gene set with targeted therapeutics (light blue in Fig. S2B) was observed. A similar association was found for the ESTROGEN_RESPONSE_EARLY gene set, likely due to the close mutual interaction between estrogen and EGFR signaling (32).

**Figure 2.**
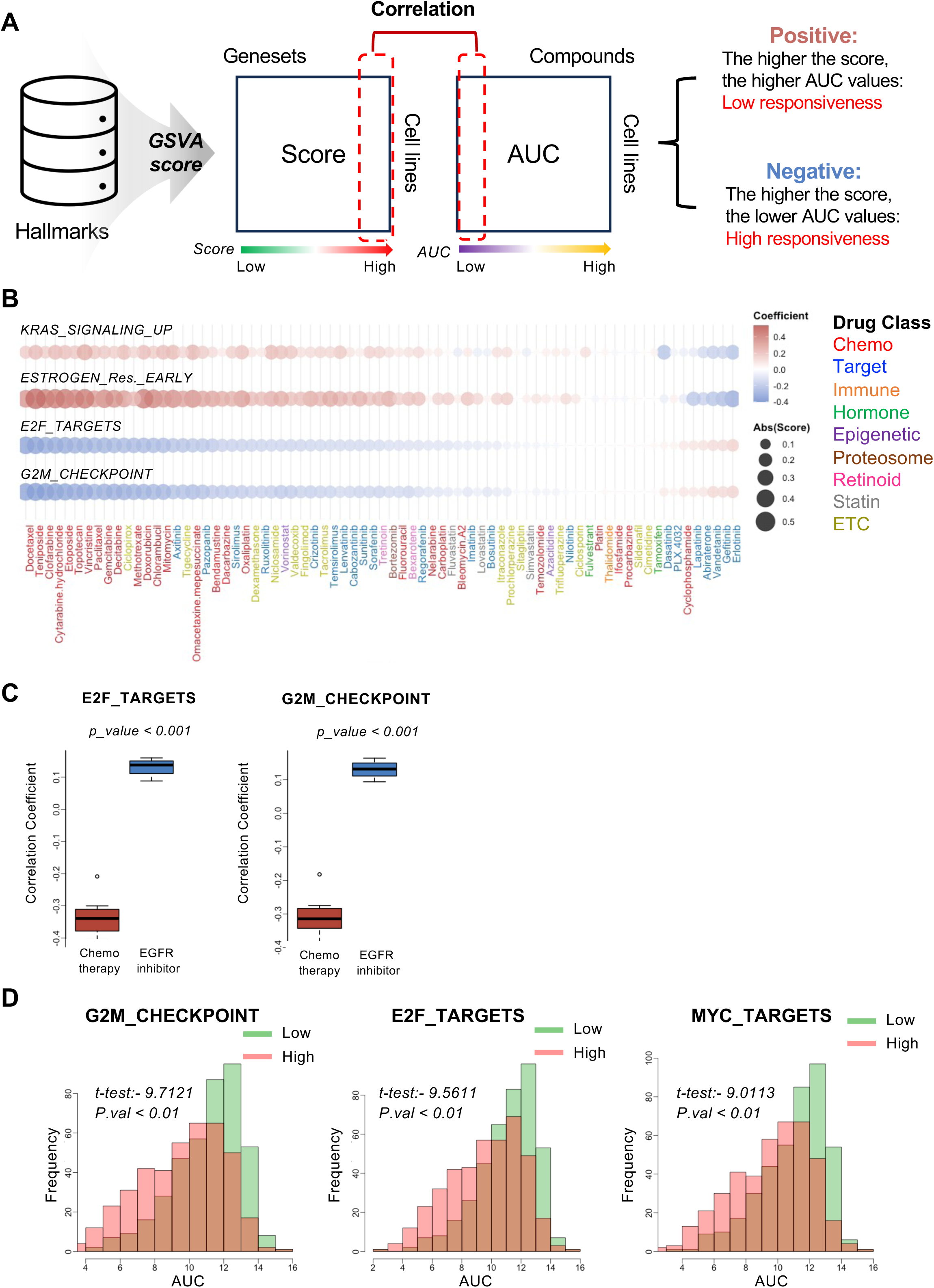
Correlation of mitosis geneset scores to chemo-responsiveness in cell line data. (A) Correlation analysis between Hallmark gene set enrichment scores and drug responsiveness across various cell lines. (B) Dot plot displaying correlation coefficients for each drug in relation to four specific Hallmark gene sets. (C) Distribution of E2F_TARGETS and G2M_CHECKPOINT gene set scores across cell lines treated with chemotherapy and EGFR inhibitors. (D) Differences in area under the curve (AUC) distributions for 14 chemotherapeutic agents, categorized by high and low scores of gene sets (E2F_TARGETS, G2M_CHECKPOINT, and MYC_TARGETS). Welch’s t-test was performed to assess the statistical significance of these differences.

These findings underscore that the high efficacy of chemotherapy is associated with active proliferation. The analysis identified a total of 14 drugs with negative correlations, all classified as chemotherapeutic agents, while 4 drugs with positive correlations were identified as EGFR inhibitors (Fig. S2B). Notably, the contrasting average correlation coefficient scores of 14 chemotherapeutics and 4 EGFR inhibitors (Fig. S2B) toward ‘E2F_TARGETS’ and ‘G2M_CHECKPOINT’ (Fig. 2C) underscore the predictive power of gene signatures associated with an active cell cycle in determining responsiveness to traditional chemotherapeutic agents. To further substantiate these findings, we focused on three typical cell cycle associated genesets: ‘G2M_CHECKPOINT’, ‘E2F_TARGETS’, and ‘MYC_TARGETS’. Cell lines were stratified into high and low score groups based on the median score of these genesets. Comparison of AUC values for 14 common chemotherapeutic agents, including paclitaxel, docetaxel, and doxorubicin, were determined in each cell cycle associated geneset. The significance of the difference in AUC values between the high-score group (red) and the low-score group (green) for each geneset was highlighted by the t-test value (Fig. 2D). Consistently, the absolute value of the t-test for each geneset reflects the magnitude of the difference, with larger values indicating a greater distinction between the groups (Fig. S2C). As shown in Figure 2D, high-score group of each geneset, indicating active cell cycle, revealed significantly lower AUC values (p < 0.001, two-sided t-test), indicating that cancer cell lines with active cell cycle gene signatures would be more sensitive to chemotherapeutics.

### Evaluation of AMSES as an *in-silico* mitotic marker for histopathological mitotic index

The strong correlation between genesets associated with an ‘active cell cycle’ and chemo-responsiveness observed in Figure 2 prompted us to evaluate AMSES, a previously defined geneset designed to capture mitotic activity in cancer cell lines (33) (Fig. 3A), as a potential therapeutic marker for predicting chemotherapeutic responsiveness in cancer patients. To validate AMSES as an *in silico* mitotic marker in cancer patients, we conducted a histopathological assessment of the mitotic index in 69 cancer patient samples. The results demonstrated a strong correlation between AMSES scores and the actual mitotic index (R² = 0.64, Pearson correlation) (Fig. 3B). Compared to cell cycle-associated gene sets, such as ‘G2M_CHECKPOINT’ (R² = 0.63), ‘E2F_TARGETS’ (R² = 0.55), and ‘MYC_TARGETS’ (R² = 0.33), the AMSES score showed a higher correlation with the actual mitotic index, supporting its utility as a molecular indicator of mitotic activity (Fig. S3A).

**Figure 3.**
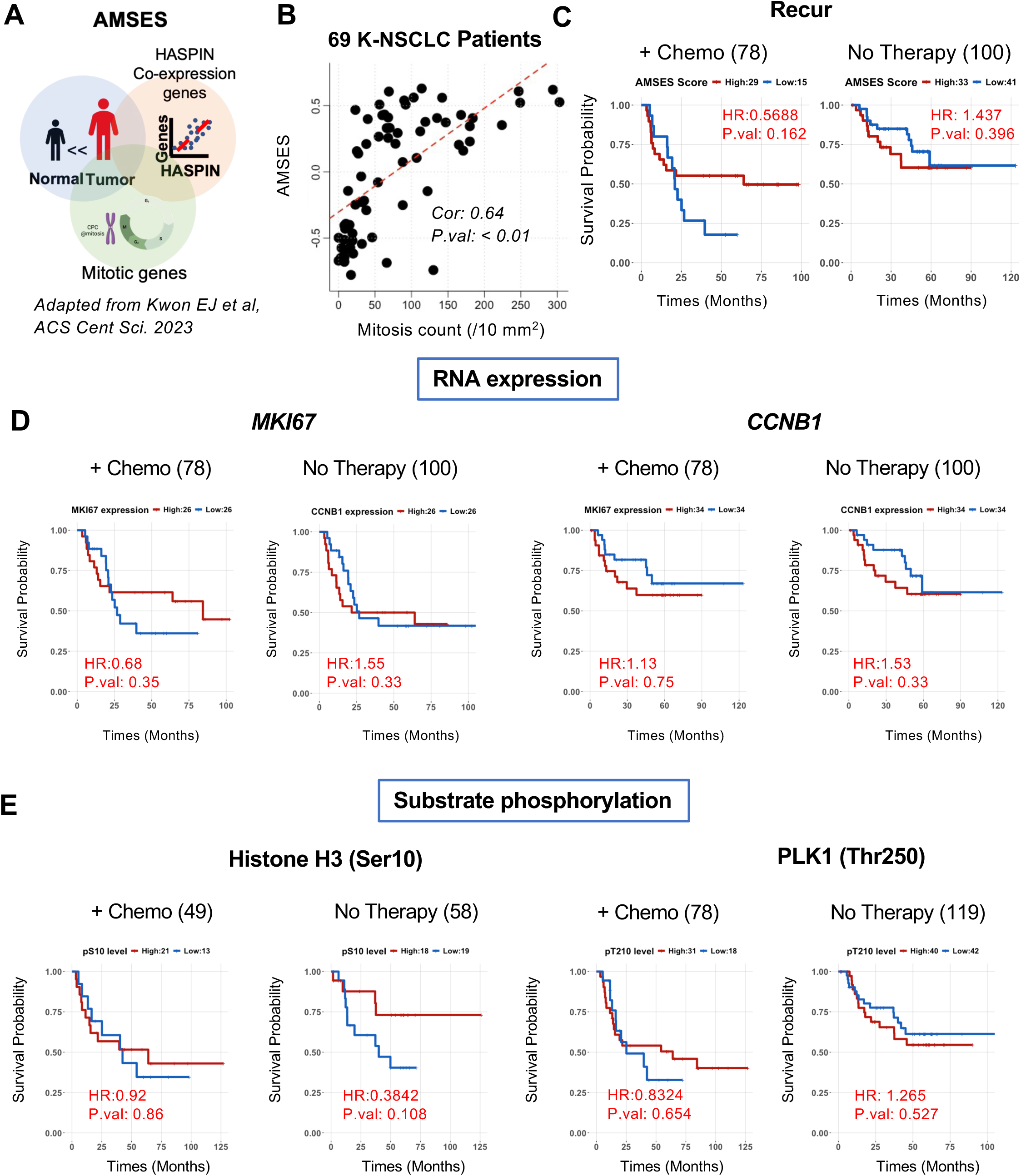
Validation of AMSES and Related Biomarkers for Predicting Recurrence Risk. (A) Composition of the Active Mitosis Gene Set Enrichment Score (AMSES), as defined in previous studies. (B) Correlation analysis between patients’ AMSES scores and their actual mitotic index values. (C) Recurrence risk analysis based on high and low AMSES scores in the “+Chemo” and “No Therapy” groups. (D) Recurrence risk analysis stratified by high and low expression levels of MKI67 and CCNB1 in the “+Chemo” and “No Therapy” groups. (E) Recurrence risk analysis based on phosphorylation levels of Histone H3 Ser10 and PLK1 T210 in the “+Chemo” and “No Therapy” groups.

However, contrary to expectations, AMSES scores failed to significantly predict recurrence risk in the group of chemotherapy, although a trend was observed (i.e., HR of high AMSES score = 0.5688 at ‘+ Chemo’, Fig. 3C). Interestingly, the expression levels of individual genes closely associated with an active cell cycle, such as *MKI67* (34) and *CCNB1* (35) (Fig. S3B), also failed to predict clinical outcomes (i.e., significant HR reduction in high group) following chemotherapy (Fig. 3D). In turn, we examined the phosphorylation status of phosphorylation levels of established mitotic markers, including Histone H3 (serine 10) (36,37) and PLK1 (threonine 210) (38,39) (Fig. S3) from phosphoproteome datasets of K-NSCLC cohort, one of unique datasets (28). Unexpectedly, these typical mitotic phosphorylation levels also failed to represent the better clinical outcome following chemotherapy (Fig. 3E). Although the groups with better clinical outcomes by chemotherapy exhibit the biomarker for active cell cycle, the individual marker for mitotic activity (or cell cycle) would be insufficient to significantly predict the chemo-responsiveness.

### Optimization of genesets for prediction of chemo-responsiveness

Instead of AMSES, consisting of 126 genes, which was able to show a desirable trend, in turn, we aimed to optimize the genes to significantly represent the clinical outcomes after chemotherapy. To this end, systematic refinement with multiple distinct methodological approaches was performed to set either ‘MIV’ or ‘recurrence risk’ as a readout (Fig. 4A). Initially we evaluated three distinct optimization approaches to derive genesets that could effectively reflect mitotic index values: i) Correlation-based analysis (Correlation), ii) Random Forest (RF), and iii) Ridge regularization logistic regression (Ridge). For each method, we constructed genesets incrementally by adding genes in descending order of their respective ranking parameters – i) Correlation coefficients, ii) Random Forest importance features, and iii) Ridge regression coefficients. For each optimization approach, we calculated 126 constructed geneset scores for patient samples and stratified them into “High” and “Low” groups using the upper and lower 33rd percentiles as thresholds (Fig. 4B). Subsequently, the hazard ratio for recurrence risk between High and Low groups among patients who received chemotherapy at the corresponding number of genes identified by i) Correlation ii) RF and iii) Ridge (Fig. 4C). The hazard ratio, indicating the risk of recurrence after chemotherapy of optimal 7 gene signature from Correlation was 0.42, the lowest hazard ratio among three approaches. These results indicate that the gene signature with 7 genes from 126 AMSES genes would predict the clinical outcome (determined by ‘recurrence risk’) effectively compared to other approaches. Validation involved assessing correlations between each method’s scores and actual mitotic counts from cancer patients as described in Figure 3B. All approaches, including AMSES, exhibited similarly high correlation coefficients (Fig. 4D) and classification accuracies in predicting mitotic activity (Fig. 4E). Additionally, AUC-ROC values, determined using the MIV, underscored the robust predictive performance of all methods as summarized in Figure 4F. Subsequently, clinical outcomes in the K-NSCLC cohorts were evaluated based on gene scores from the optimized genesets. Notably, patients in the high-score group from the Correlation approach demonstrated a significantly reduced risk of recurrence among the 78 patients who received chemotherapy (p = 0.02), an effect not observed in the 100 patients without chemotherapy (Fig. 4G). However, statistical significance was not observed in the other approaches (Fig. 4G)

**Figure 4.**
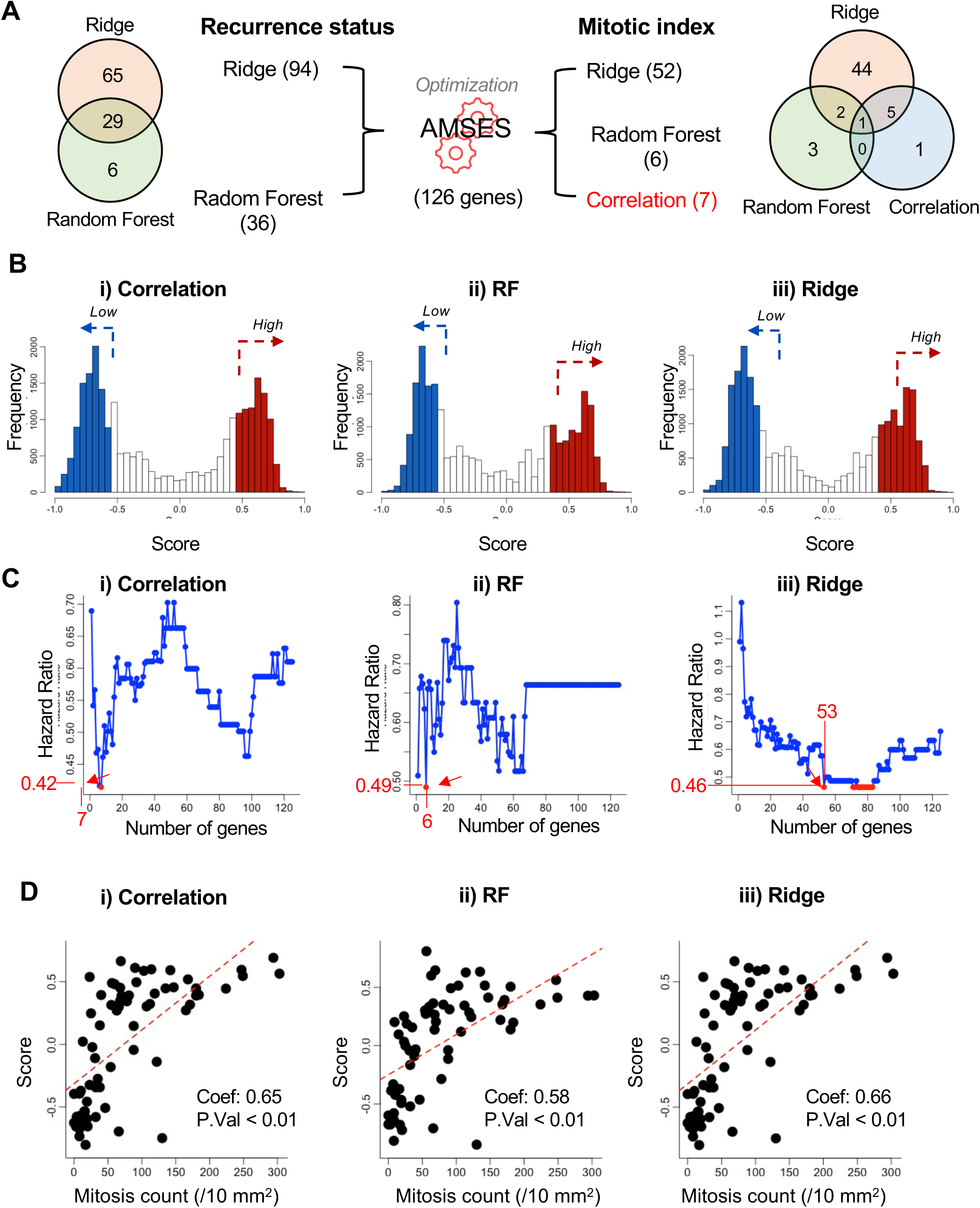
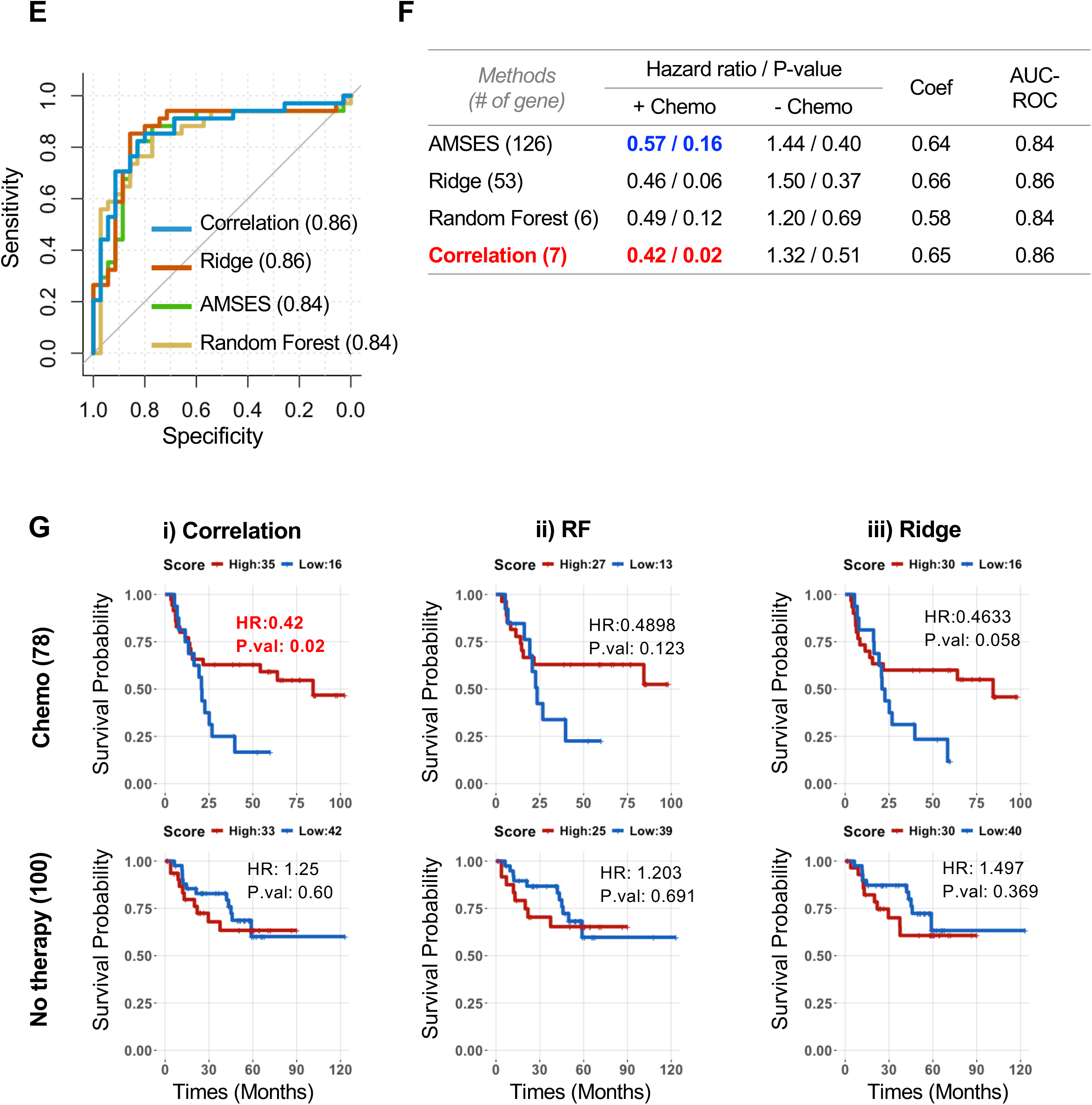
Optimization and Validation of AMSES Gene Set for Recurrence Prediction. (A) Optimization of the AMSES gene set based on Mitotic Index and Recurrence Status. Overlapping genes among optimized gene sets were illustrated using a Venn diagram. (B) Distribution of optimized gene set scores among patient groups, categorized into tertiles: High (top 33%) and Low (bottom 33%). (C) Recurrence hazard ratios for high versus low groups of gene sets based on the number of genes included. (D) Correlation analysis between patients’ optimized gene set scores (Correlation, RF, Ridge) and their actual Mitotic Index values. (E) ROC curve displaying true-positive versus false-positive rates for recurrence status prediction. (F) Summary table comparing AMSES and optimized gene sets (Correlation, RF, Ridge) in terms of recurrence hazard ratio, p-value, Pearson correlation with actual Mitotic Index values, and AUC-ROC scores from panel E. (G) Risk analysis of recurrence for patients with low and high optimized gene set scores in the “+Chemo” (upper panels) and “No Therapy” (lower panels) groups.

Additionally, to specifically optimize for ‘recurrence risk,’ we also evaluated two distinct approaches: (i) Random Forest and (ii) Ridge regularization logistic regression. Similar to the MIV optimization process, genes were incrementally added based on their ranking parameters, and gene set scores were calculated for patient samples, with the top and bottom 33% defining “High” and “Low” risk groups (Fig. S4A and B, left panels). The hazard ratios between these groups in chemotherapy-treated patients were then assessed (Fig. S4A and B, middle panels). The optimal gene sets identified contained 36 genes for RF and 94 genes for Ridge, both yielding a hazard ratio of 0.52 but without statistical significance (Fig. S4A and B, right panels).

Collectively, these results demonstrated that AMSES genes were more strongly associated with ‘mitotic activity’ (Fig. S4C) compared to ‘recurrence risk’ (Fig. S4D). This conclusion was further supported by the evaluation of metrics such as accuracy, precision, recall, F1-score, and AUC-ROC (Fig. S4E). Among the three methods evaluated, the correlation-based approach demonstrated the most robust predictive performance for clinical outcomes, particularly in assessing recurrence risk. Based on these findings, we designated the optimized geneset derived from the correlation-based approach as ‘AMSES for chemo-responsiveness’ (A4CR).

### Validation of A4CR for Predicting Chemotherapy Response Across Multiple Datasets

To evaluate broader applicability, the A4CR (Fig. 4) was validated using independent cell line response datasets and patient datasets from TCGA that included chemotherapy and patient survival data. The predictive value of A4CR was further validated using three independent cancer cell line drug response datasets such as CTRP (40), PRISM (41) and GDSC (42). Cell lines with higher A4CR scores demonstrated significantly lower AUC values (average t-test values = −5.77, p < 0.01), indicating increased sensitivity to chemotherapeutic agents (Fig. 5A). In contrast, the genesets derived from “RF,” “Ridge,” or the original “AMSES” demonstrated less effective predictive capability for chemotherapy sensitivity across all three datasets, CTRP (Fig. S4A), GDSC (Fig. S5B) and PRISM (Fig. S5C) compared to A4CR as summarized in Figure S5D. We further assessed the predictability of A4CR using publicly available clinical data. Among 830 lung cancer patients (both LUAD and LUSC) in the TCGA dataset, a total of 302 patients who received chemotherapy (+ Chemotherapy) were categorized into two groups: 231 patients who received adjuvant chemotherapy and 43 patients who received chemotherapy after recurrence. Detailed subgroup classifications of these patients are provided (Fig. S5E). The predictability of A4CR was assessed by comparing the survival rate of 191 patients with adjuvant chemotherapy (Adjuvant Chemo)—excluding 38 patients with missing data and new primary tumor (N.PT) incidence—to 404 patients who received no adjuvant therapy (No Adjuvant)—excluding 124 patients with missing data and N.PT (Fig. 5B). This analysis aimed to determine the association between A4CR and chemotherapy response in a well-defined patient cohort. Consistent with our previous findings, patients with higher A4CR scores exhibited significantly lower recurrence risk only in the adjuvant chemotherapy group but not in the ‘No Adjuvant’ group (Fig. 5C). The improved performance of A4CR for survival prediction in the adjuvant chemotherapy group, as reflected by lower hazard ratio (HR) and p-value, further supports its potential as a predictive biomarker for chemotherapy response compared to RF and Ridge gene sets (Fig. S5F). Additionally, a greater proportion of patients with high A4CR scores in the adjuvant chemotherapy group achieved favorable drug responses, such as Complete Response (CR) or Partial Response (PR) (Fig. 5D). However, when we extended the analysis to other TCGA cancer cohorts, including breast cancer (BRCA), colon adenocarcinoma (COAD), and head and neck squamous cell carcinoma (HNSC), the number of patients who experienced recurrence after receiving adjuvant chemotherapy was too small for meaningful statistical analysis (n = 9 for BRCA, n = 0 for COAD, and n = 3 for HNSC). This limitation highlights the need for further validation in larger, independent cohorts to determine the broader applicability of A4CR across different cancer types.

**Figure 5.**
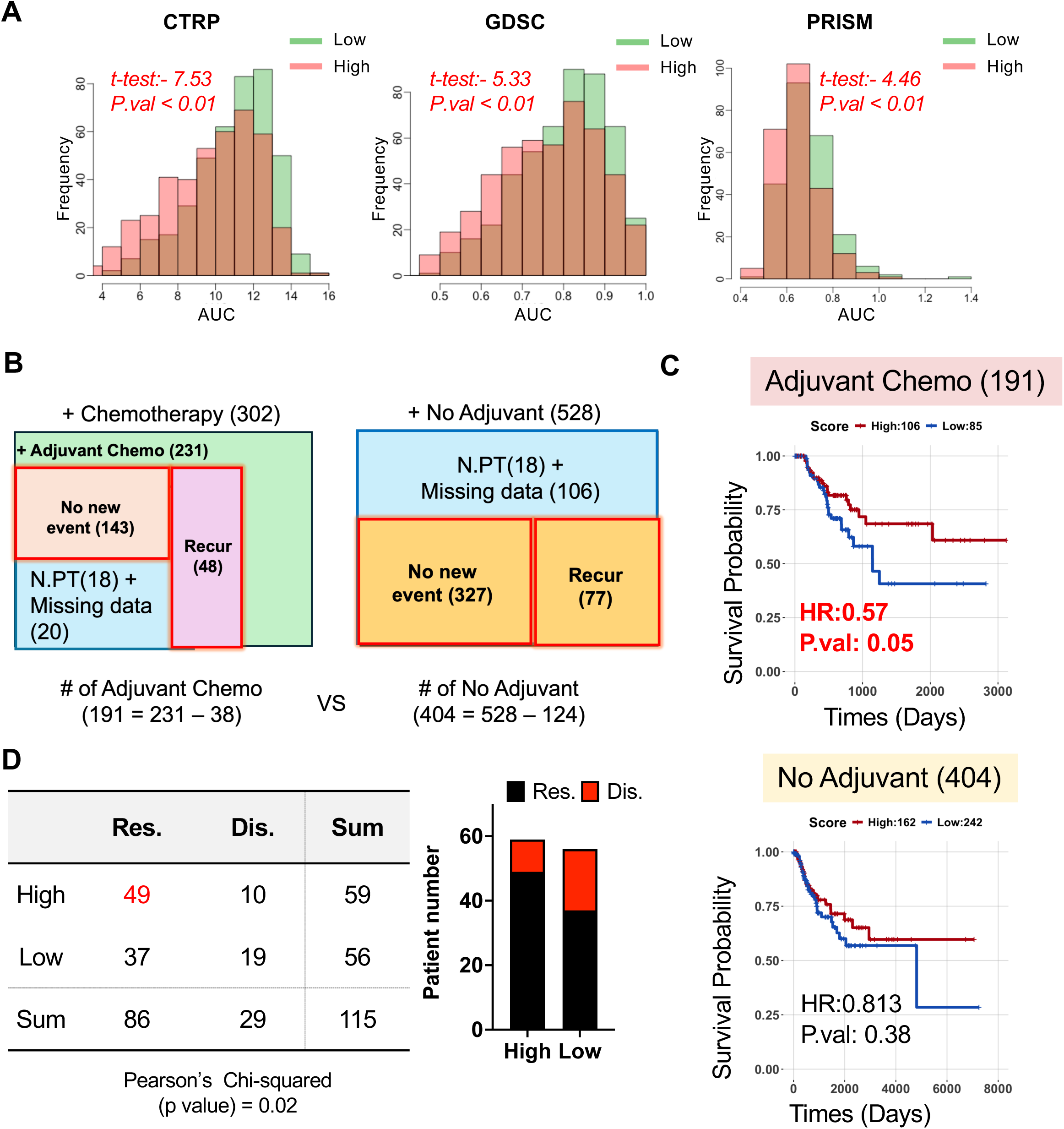
A4CR Gene Set Analysis and Its Clinical Implications. (A) Comparison of area under the curve (AUC) distributions for 14 chemotherapies, stratified by high and low A4CR gene set scores in CTRP, GDSC and PRISM datasets. (B) Recurrence risk analysis based on A4CR scores in the “Adjuvant Chemo” and “No Adjuvant” groups. (C) Contingency table and corresponding p-value evaluating the association between A4CR scores and chemotherapy responsiveness.

## Discussion

Chemotherapy, despite its widespread use in cancer treatment, lacks reliable biomarkers for predicting treatment response, unlike targeted or immune therapies that rely on specific therapeutic biomarkers for patient stratification. Multiple studies have attempted to identify chemotherapy response biomarkers, investigating DNA repair pathways, cell cycle regulators, and tumor microenvironment factors (43–45). However, a standardized and clinically applicable biomarker panel remains elusive. The efficacy of chemotherapy is fundamentally linked to cellular proliferation, as these agents primarily target actively dividing cells. Numerous clinical studies have demonstrated a strong correlation between the mitotic index—an indicator of active mitosis—and clinical outcomes following chemotherapy (46–48). For systemic demonstration, we took advantage of the K-NSCLC cohort including phosphoproteome datasets linked to clinical outcomes, encompassing therapy regimens and recurrence data, alongside transcriptome datasets (28). This comprehensive data enables the evaluation of mitotic activity using phosphoproteins such as Histone H3 and PLK1 in relation to chemotherapy outcomes. Unexpectedly, mitosis-specific phosphoproteins like Histone H3 (Ser10) failed to significantly predict clinical outcomes after chemotherapy (Fig. 3E), while the mitotic index value (MIV) demonstrated strong predictive value for both recurrence and survival in the K-NSCLC cohort (Figs. 1D and E). Similarly, the AMSES geneset, containing 126 mitosis-associated genes, showed only limited predictive power (Figs. 3B and C). To enhance the utility of transcriptome signature for chemo-responsiveness, the AMSES geneset was optimized using machine learning approaches, including correlation analysis, Random Forest, and Ridge regression models (Fig. 4). Among these methods, the correlation-based approach showed superior performance, likely due to its focus on genes with strong linear associations with mitotic activity, effectively capturing the biological link between cell proliferation and chemotherapy sensitivity. In contrast, the Ridge and Random Forest methods introduced complexity, potentially diluting the predictive strength by modeling non-linear relationships or penalizing coefficients (Fig. 4).

In parallel with advancements in machine learning and sequencing technologies, various algorithms have been developed to predict chemotherapy responsiveness (49–52). While previous studies have utilized diverse data types such as CT imaging, genomic profiles, and pathological data, in combination with machine learning approaches, to develop predictive models and identify factors associated with chemotherapy responsiveness across various cancer types(49,50,53), our study distinguishes itself by adopting a more targeted approach. We began with the straightforward notion of determining chemotherapy responsiveness through the concept of ‘active mitosis or active cell cycle,’ as demonstrated by the clinical outcomes directly associated with the actual mitotic index of individual cancer patients in the K-NSCLC cohort (Fig. 1). The superior predictability of A4CR in lung cancer patients (Fig. 5) stems from the systematic optimization of AMSES genes using machine learning techniques. By focusing on genes associated with mitotic activity, A4CR captures the proliferative status of tumors, a critical determinant of chemotherapy response. A4CR serves as a practical tool for predicting chemotherapy responsiveness, aiding clinicians in identifying patients who may not benefit from chemotherapy, reducing unnecessary side effects while maximizing the potential for personalized treatment and improved patient outcomes. Furthermore, A4CR’s approach can be expanded to develop biomarkers tailored to specific chemotherapy regimens, guiding more effective and individualized cancer therapies. For instance, the MD Anderson chemotherapy response calculator predicts disease-free survival after preoperative anthracycline-based chemotherapy based on histological type, grade, and tumor size in breast cancer. Similarly, A4CR can be proposed as a transcriptome-based predictive tool for chemotherapy responsiveness, with potential applications across various tumor types.

In summary, our study highlights the critical role of tumor cell proliferation, as measured by the mitotic index, in predicting chemotherapy responsiveness. By combining this biological insight with machine learning optimization, A4CR represents a significant step toward enhancing the precision of chemotherapy predictions in clinical settings, ultimately contributing to more personalized and effective cancer care.

## Author Contributions

HJ.C conceived the overall study design and led the experiments. EJ.K primarily conducted the experiments and data analysis and contributed to critical discussions of the results. HS.H obtained the mitotic index from cancer patients and provided clinical insights. E.C and JY.A contributed to in silico data acquisition. All authors have reviewed and approved he final version of the manuscript.

## Data Availability

All data produced in the present study are available upon reasonable request to the authors

## Acknowledgements

This work was supported by the ABC-based Regenerative BioTherapeutics (ABC project) grant (RS-2024-00432867) and ERC grant (RS-2023-00218543) funded by the Korea government (the Ministry of Health & Welfare)

---

**Figure S1.**
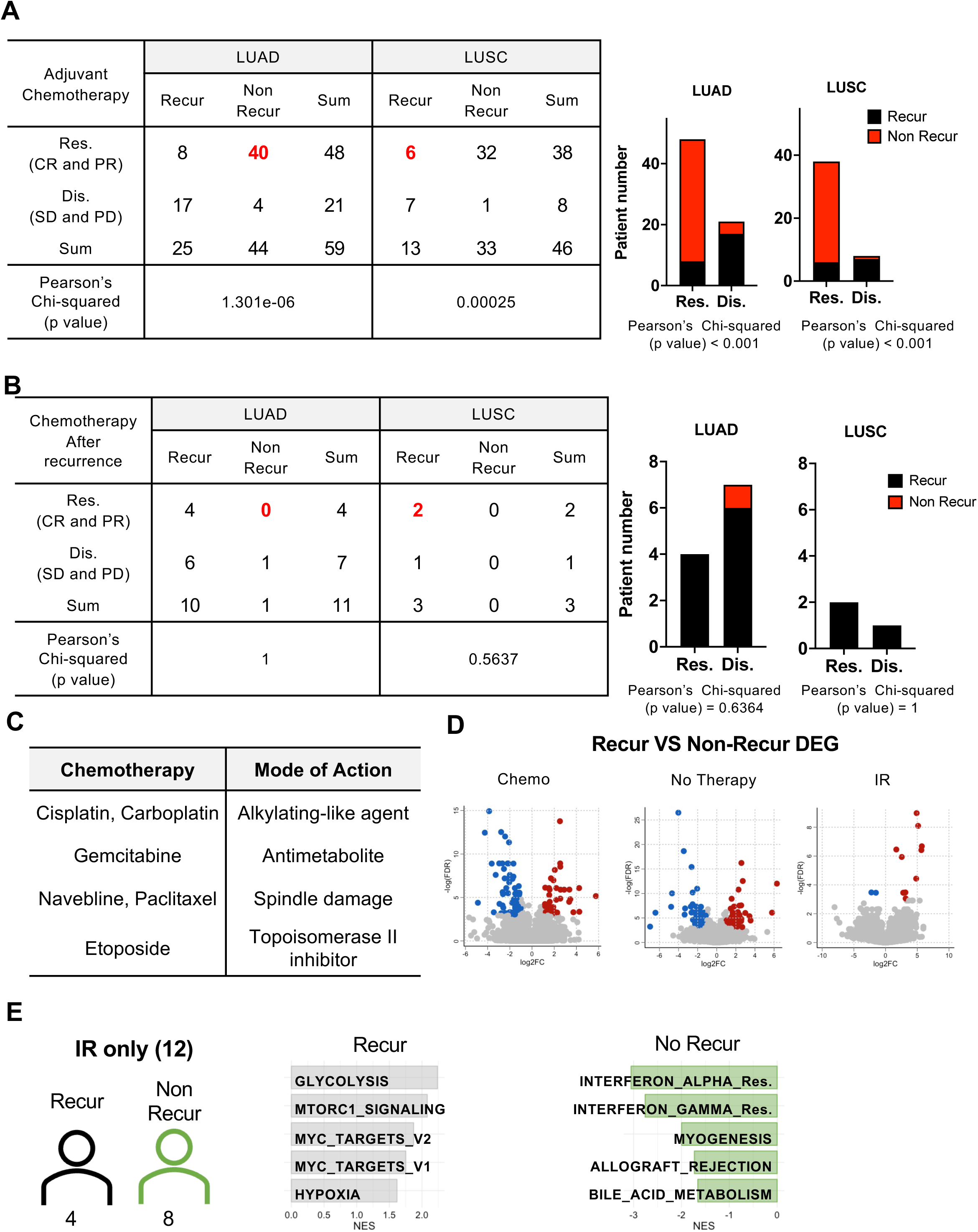

**Figure S2.**
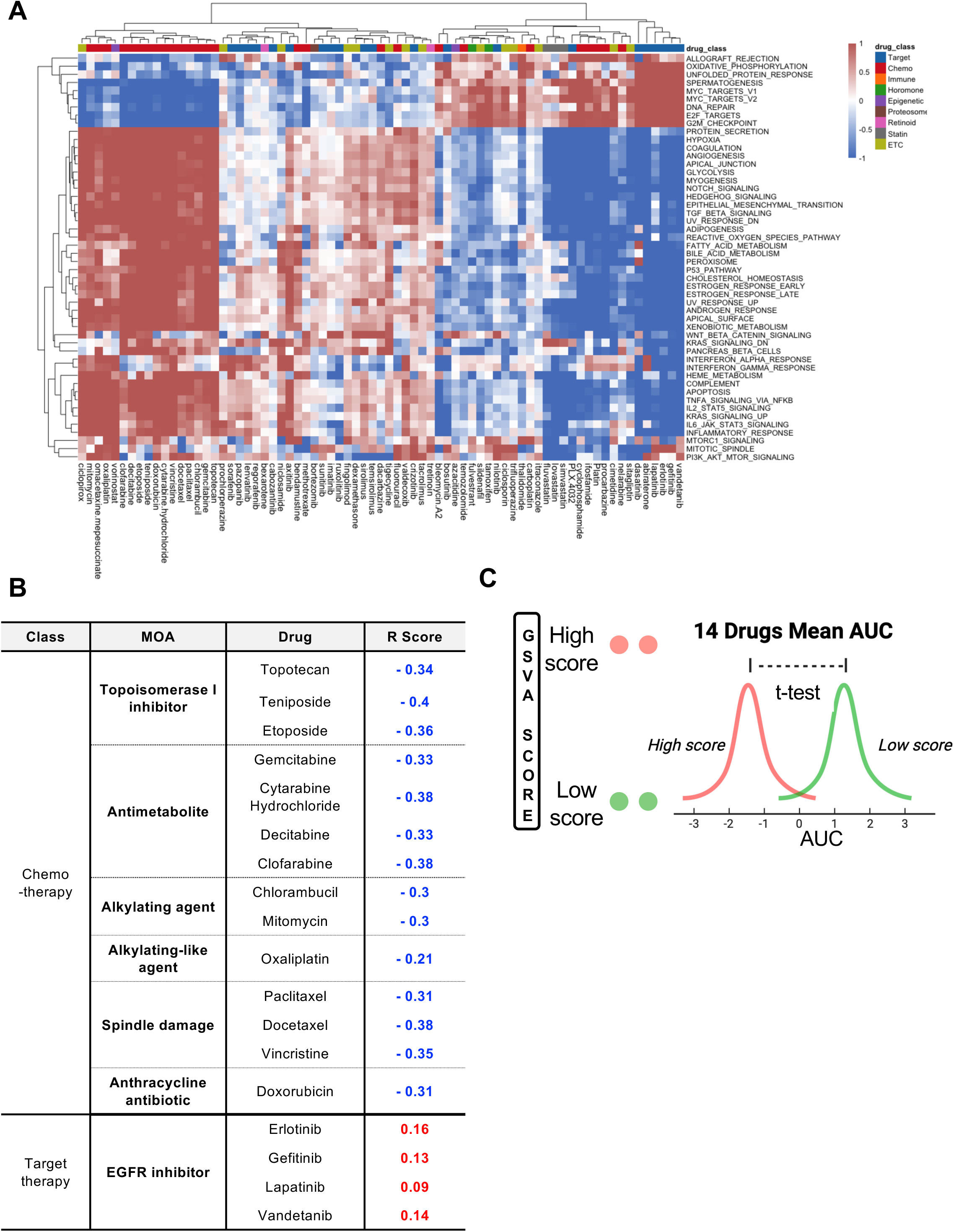

**Figure S3.**
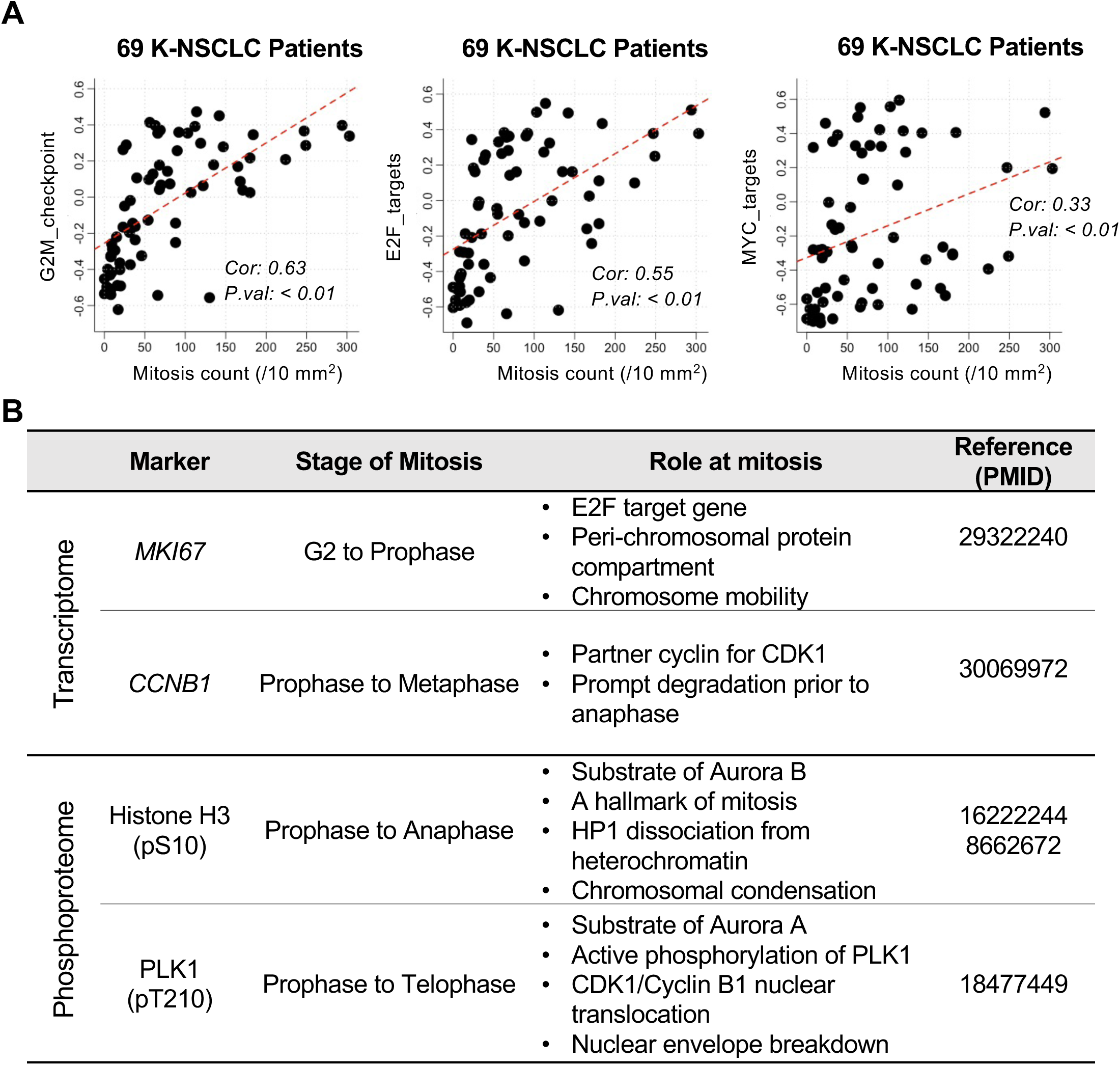

**Figure S4.**
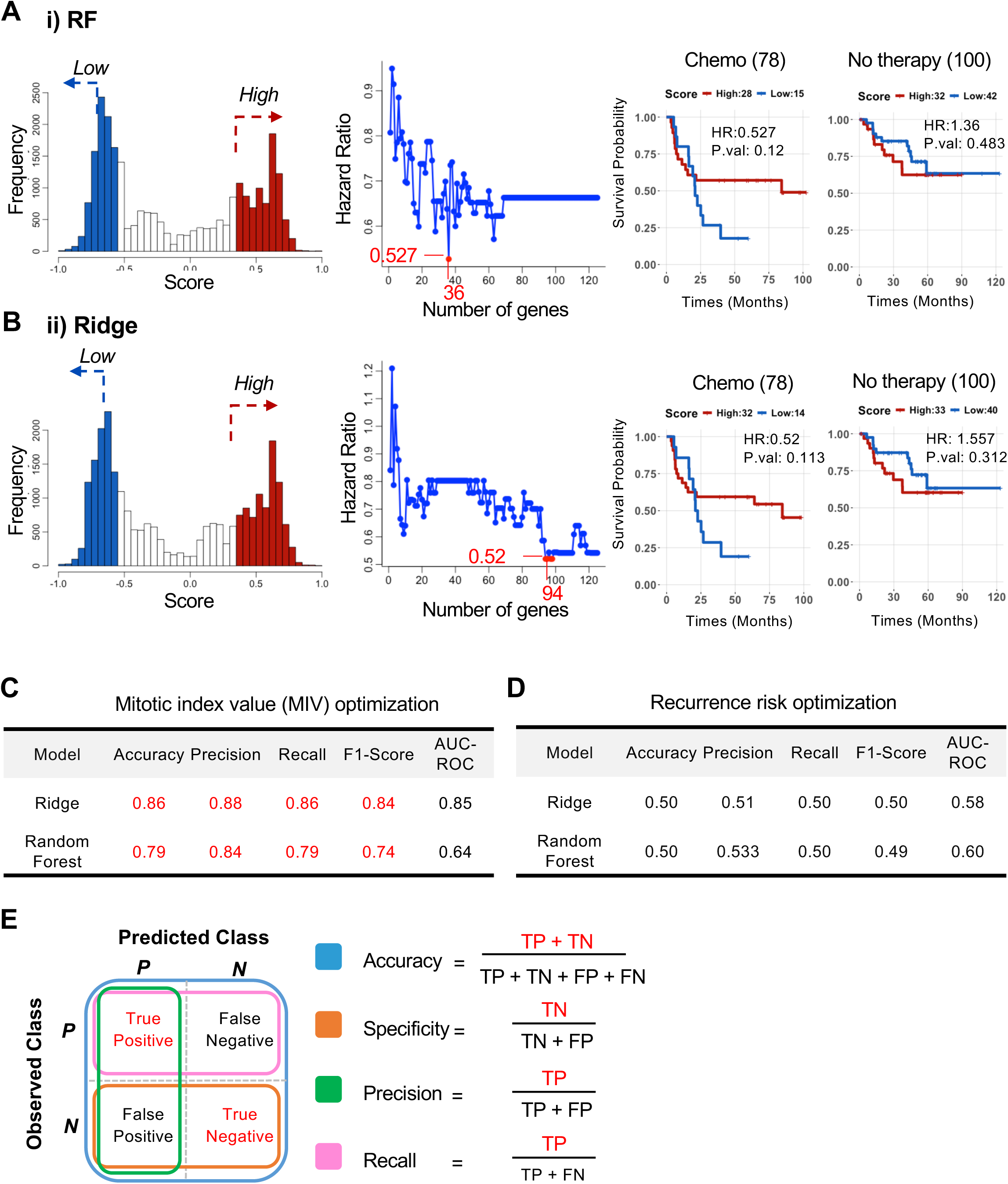

**Figure S5.**
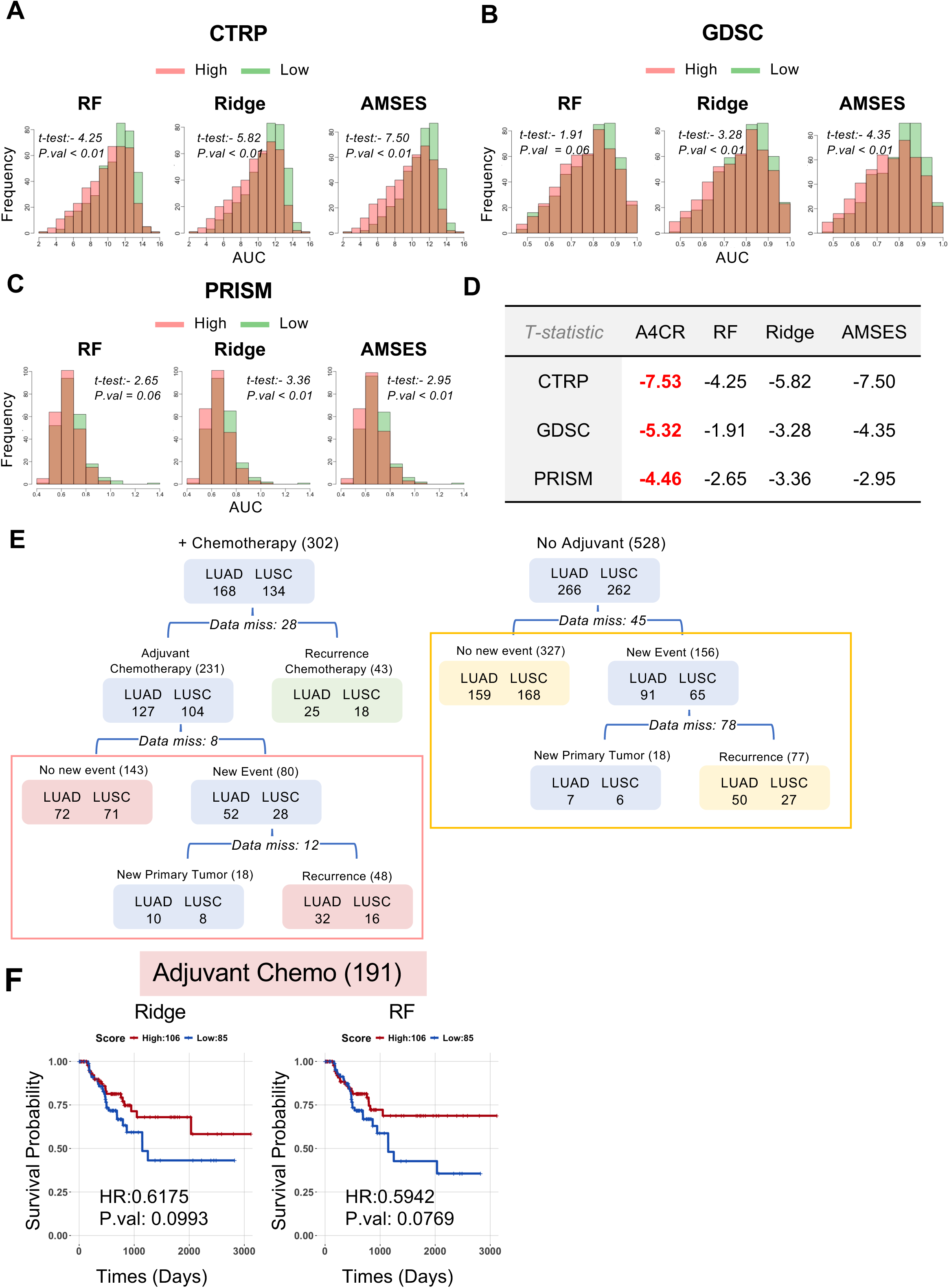

